# Quantitative measurement of IgG to SARS-CoV-2 proteins using ImmunoCAP

**DOI:** 10.1101/2020.11.09.20228411

**Authors:** Behnam Keshavarz, Joesph R. Wiencek, Lisa J. Workman, Matthew D. Straesser, Lyndsey M. Muehling, Glenda Canderan, Fabrizio Drago, Catherine A. Bonham, Jeffrey M. Sturek, Chintan Ramani, Coleen A. McNamara, Judith A. Woodfolk, Alexandra Kadl, Thomas A.E. Platts-Mills, Jeffrey M. Wilson

**Author notes:** Corresponding Author: Jeffrey M. Wilson, MD, PhD Department, Division of Allergy & Clinical Immunology, University of Virginia, P.O. Box 801355, Charlottesville, VA, 22908-1355, USA, Thomas A.E. Platts-Mills, MD, PhD, FRS, Division of Allergy & Clinical Immunology, University of Virginia, P.O. Box 801355, Charlottesville, VA, 22908-1355, USA. Department of Pathology, Immunology, and Microbiology. Vanderbilt University School of Medicine, Nashville, TN.

## Abstract

**Background:** Detailed understanding of the immune response to SARS-CoV-2, the cause of coronavirus disease 2019 (COVID-19), has been hampered by a lack of quantitative antibody assays.

**Objective:** To develop a quantitative assay for IgG to SARS-CoV-2 proteins that could readily be implemented in clinical and research laboratories.

**Methods:** The biotin-streptavidin technique was used to conjugate SARS-CoV-2 spike receptor-binding-domain (RBD) or nucleocapsid protein to the solid-phase of the ImmunoCAP resin. Plasma and serum samples from patients with COVID-19 (n=51) and samples from donors banked prior to the emergence of COVID-19 (n=109) were used in the assay. SARS-CoV-2 IgG levels were followed longitudinally in a subset of samples and were related to total IgG and IgG to reference antigens using an ImmunoCAP 250 platform.

**Results:** Performance characteristics demonstrated 100% sensitivity and 99% specificity at a cut-off level of 2.5 µg/mL for both SARS-CoV-2 proteins. Among 36 patients evaluated in a post-hospital follow-up clinic, median levels of IgG to spike-RBD and nucleocapsid were 34.7 µg/mL (IQR 18-52) and 24.5 µg/mL (IQR 9-59), respectively. Among 17 patients with longitudinal samples there was a wide variation in the magnitude of IgG responses, but generally the response to spike-RBD and to nucleocapsid occurred in parallel, with peak levels approaching 100 µg/mL, or 1% of total IgG.

**Conclusions:** We have described a quantitative assay to measure IgG to SARS-CoV-2 that could be used in clinical and research laboratories and implemented at scale. The assay can easily be adapted to measure IgG to novel antigens, has good performance characteristics and a read-out in standardized units.

## Introduction

Serological assays that quantify antibodies specific for severe acute respiratory syndrome coronavirus (SARS-CoV)-2 represent an important tool in the investigation of the epidemiology and immunology of this novel coronavirus, the cause of the coronavirus disease (COVID-19) pandemic [1-3]. Many studies have reported on antibodies specific for SARS-CoV-2, however most of the assays used in these studies have had qualitative and/or semi-quantitative read-outs and very few have used standardized units (eg, µg/mL) [4-12]. The ImmunoCAP assay developed by Phadia/Thermo Fisher is often considered the gold-standard for quantitative detection of IgE antibodies to allergens in both clinical and research laboratories. The platform was designed for IgE detection but can also be used to quantify other antibody isotypes/subclasses including IgG and IgG4 [13, 14]. The assay has high accuracy, consistency and reproducibility and has a read-out in standardized units (ie, IU/mL for IgE and μg/mL for IgG and IgG4) [15]. A major strength of the ImmunoCAP is that antibodies specific for multiple antigenic targets can readily be assessed in parallel. With the emergence of the coronavirus pandemic we asked whether the ImmunCAP could be used to measure IgG to SARS-CoV-2 proteins. Here we describe a novel assay that takes advantage of the biotin-streptavidin technique to link commercially available SARS-CoV-2 proteins to the high-capacity absorbent solid-phase of the ImmunoCAP [16, 17]. The assay was developed using plasma and serum samples from established COVID-19 patients and also samples from control subjects banked prior to the pandemic. Our results suggest that the ImmunoCAP-based approach has good performance characteristics and offers several advantages over many commercial assays currently in use.

## Materials and Methods

### Reagents

Recombinant SARS-CoV-2 S1 receptor-binding domain subunit (RBD) and recombinant SARS-CoV-2 nucleocapsid protein which had been expressed in HEK293 cells were purchased from RayBiotech (Peachtree Corners, GA). Human coronavirus (HCoV-229E) spike protein (S1 subunit, His Tag) and human coronavirus (HCoV-OC43) spike protein (S1 subunit, His Tag), protein were purchased from Sino Biological (Wayne, PA). The recombinant monoclonal anti-SARS-CoV-2 spike glycoprotein S1 antibody (CR3022) was purchased from Abcam (Cambridge, MA). Streptavidin ImmunoCAPs and tetanus toxoid ImmunoCAPs, as well as ImmunoCAPs to measure total IgG, were purchased from Thermo Fisher Scientific (Portage, MI). Galactose-α-1,3-galactose-β-1,4-GlcNAc-HSA (14 atom spacer) was purchased from Dextra (Reading, UK).

### COVID-19 patients and controls

De-identified plasma samples from 15 patients hospitalized with COVID-19 were available through the clinical laboratory for initial assay development carried out under the Common Rule. Paired serum and plasma samples were available from 36 patients with COVID-19 seen in outpatient post-hospitalization follow-up clinic as part of a University of Virginia (UVA) Institutional Review Board (IRB) approved investigation. Serum samples were available from 17 patients at various longitudinal time points during inpatient admission for management of COVID-19 as part of a UVA IRB-approved investigation. Of these 17 patients, 8 also had a sample available from a subsequent outpatient follow-up clinic. Serum from allergic and non-allergic adults (n=109) obtained prior to December 2019 as part of a UVA IRB-approved investigation were used as reference controls.

### IgG ImmunoCAP assays

The biotin-streptavidin technique was used to link target antigens to the solid-phase of the ImmunoCAP assay. Biotinylation was performed using EZ-Link Sulfo-NHS-LC-Biotin (ThermoScientific, Waltham, MA). Briefly, to 500 µL of 2 mg/mL protein solution, 60 µl of 2 mg/mL of biotin was added. Mixtures were placed in the dark and incubated for 4 hours at room temperature. The biotinylated proteins were dialyzed against PBS at 4°C for 24 hours with three buffer changes. In initial optimization experiments, different amounts of biotinylated spike-RBD and nucleocapsid proteins were conjugated to the solid phase (4 µg, 2 µg, 1 µg, 0.5 and 0.25 µg). Subsequent experiments used 1.25 µg of SARS-CoV-2 protein to generate each ImmunoCAP. For the α-Gal assay, 2 µg of biotinylated α-Gal-HSA was used to generate each ImmunoCAP. Total IgG was measured according to manufacturer’s instructions using a 1:200 benchtop dilution. Assays were carried out with an ImmunoCAP 250 (Thermo Fisher Scientific).

### Statistical analysis

Antibody levels were non-normally distributed and were compared by Mann-Whitney U test. The cut-off threshold of the assay as well as sensitivity and specificity of the assay for spike-RBD and nucleocapsid was determined using receiver operating characteristics (ROC) curve analysis. Correlation between concentration of the monoclonal antibody CR3022 and assay read-out were determined using simple linear regression. All analyses were performed using GraphPad Prism V8.4 (San Diego, CA).

## Results

### Assay development

The assay was initially developed using plasma from 15 de-identified patients hospitalized with COVID-19 who had been confirmed to have IgG to SARS-CoV-2 with the FDA-EUA-approved Abbott Architect i2000 immunoassay. To measure IgG antibody to SARS-CoV-2, streptavidin-linked immunosorbents were prepared using 1.25 μg of biotinylated spike-RBD or nucleocapsid protein, as determined in preliminary optimization experiments (online suppl. Fig. 1). Dilution curves carried out at 1:100, 1:500, 1:2,500, 1:12,500 and 1:62,500 (accounting for the 1:100 onboard dilution) exhibited parallelism for both SARS-CoV-2 proteins (Fig. 1A). Observed values with the nucleocapsid ImmunoCAP assay generally aligned with the results of the Abbott assay, which used nucleocapsid on the solid phase, in a head-to-head comparison (Fig. 1B). Initial results using matched plasma and serum from seroconverted patients showed negligible differences, demonstrating both types of specimens can be used in this assay (online suppl. Fig. 2). To address the quantitative accuracy of the assay we utilized a monoclonal anti-SARS-CoV-2 spike S1 IgG1 antibody (CR3022) [5]. The concentration of CR3022 mAb [5] used in the assay correlated closely with the assay read-out (R^2^=0.97, *P*<0.001) (Fig. 1C).

**Fig. 1.**
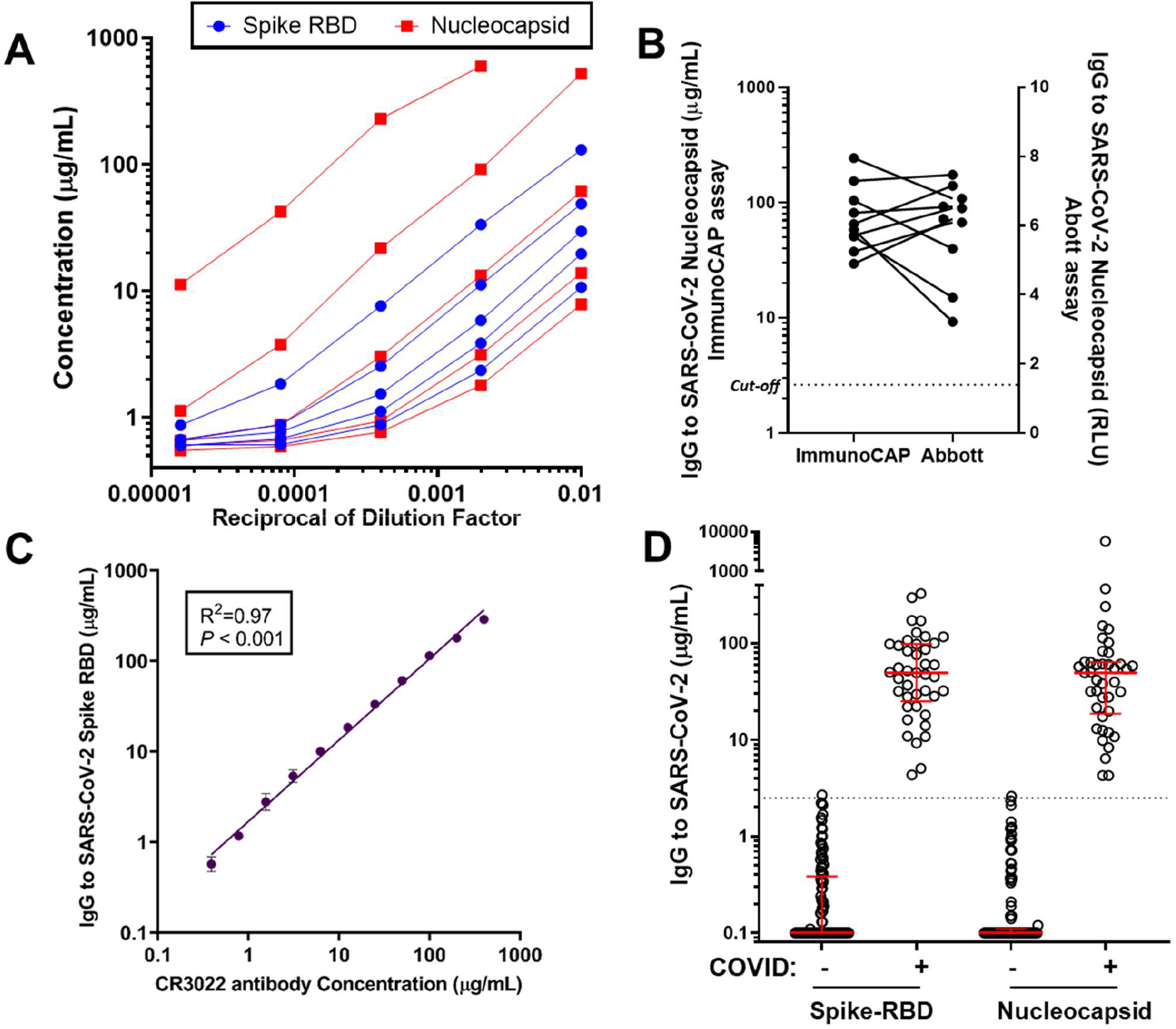
**(A)** Dilution curves for IgG to SARS-CoV-2 spike-RBD and nucleocapsid (5-fold dilutions, n=5). **(B)** Comparison of results of IgG to SARS-CoV-2 nucleocapsid using ImmunoCAP and Abbott assay (n=10). **(C)** Concentration response curve using the anti-spike-glycoprotein S1 monoclonal antibody CR3022 (2-fold dilutions carried out in triplicate ± SD). **(D)** IgG to SARS-CoV-2 spike-RBD and nucleocapsid in COVID-19 patients (n=41) and controls (n=109).

**Fig. 2.**
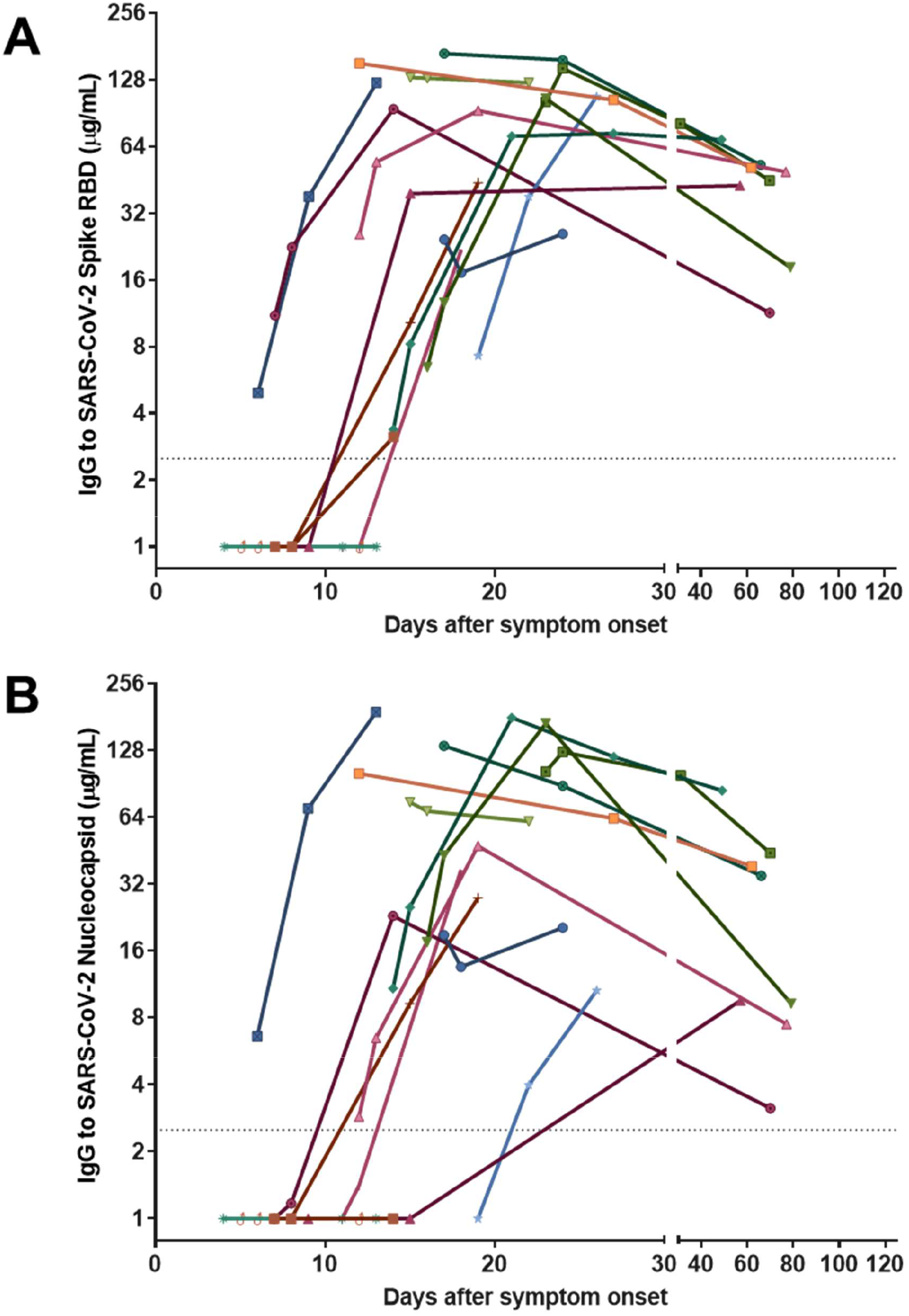
Longitudinal sampling of IgG to SARS-CoV-2 spike-RBD **(A)** and nucleocapsid **(B)** with ImmunoCAP in 17 patients hospitalized with COVID-19. All samples after day 40 were obtained from a post-hospital follow-up clinic.

To determine an optimal cut-off limit and non-specific binding in the assay, samples from COVID-19 patients (n=41) and samples from allergic and non-allergic donors banked prior to the emergence of COVID-19 (n=109) were assayed using ImmunoCAPs conjugated with SARS-CoV-2 protein and also unconjugated “naked” streptavidin ImmunoCAPs. The COVID-19 samples were obtained from the 15 hospitalized patients described above, as well as 26 patients evaluated in an outpatient follow-up clinic following hospitalization for COVID-19 [18]. IgG binding to naked streptavidin ImmunoCAPs was observed at low levels in both COVID-19 and pre-COVID-19 samples (online suppl. Fig. 3). To account for this background binding, the result obtained with the streptavidin ImmunoCAP was henceforth subtracted from the result obtained using viral antigen for each sample that was assayed. At a cut-off threshold of 2.5 µg/mL the assay had 100% sensitivity and 99% specificity in receiver operating characteristic (ROC) curve analysis for both the spike-RBD and nucleocapsid assay (Fig. 1D). Of note, this value is only marginally different from the value of 2 µg/mL which is the lower end of the assay’s measuring range according to the manufacturer’s instructions and within the range in which the assay exhibited parallelism in dilution experiments (Fig. 1A).

**Fig. 3.**
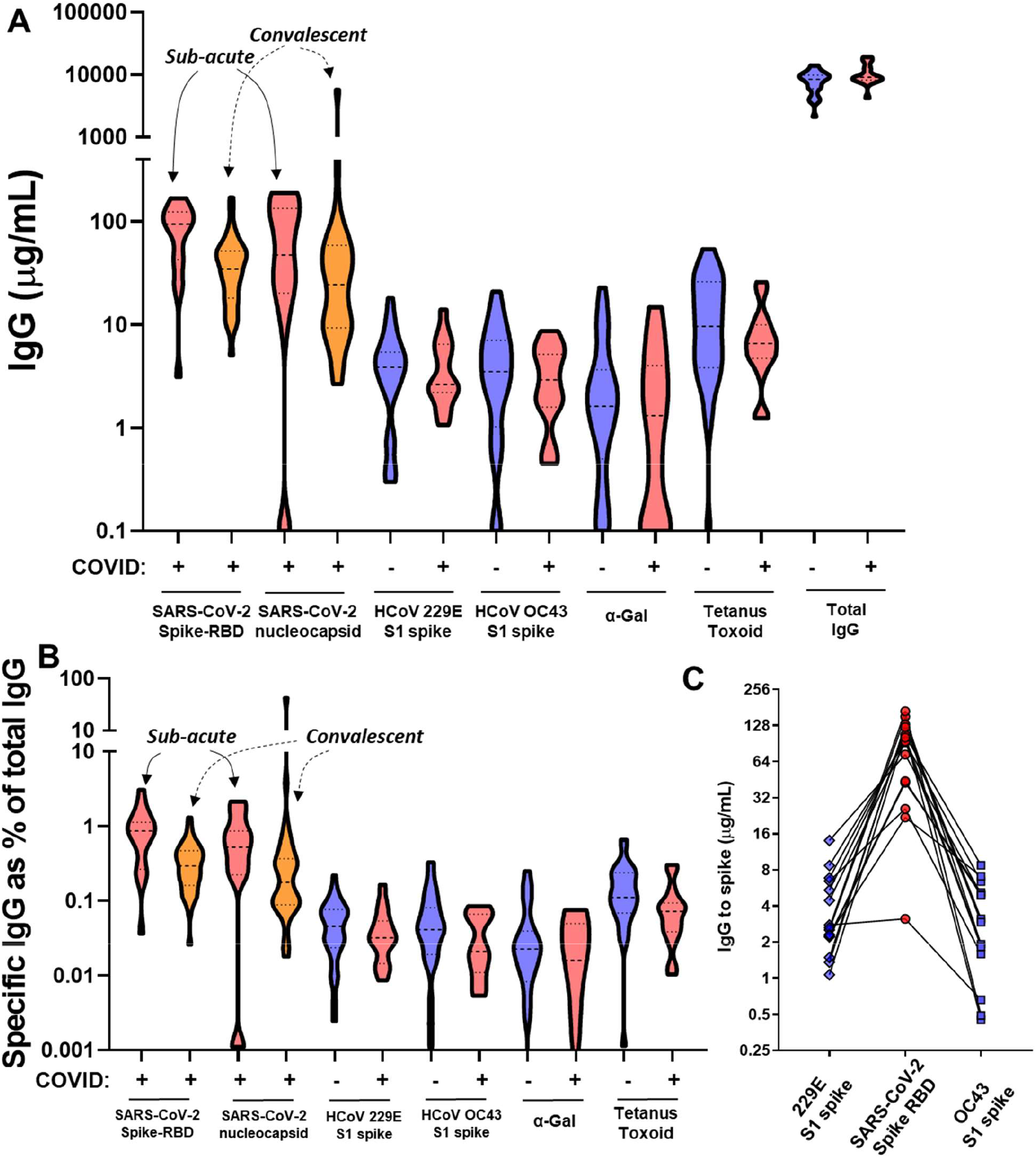
**(A)** Levels of IgG to SARS-CoV-2 spike-RBD, non-SARS coronaviruses and reference antigens with ImmunoCAP, and also total IgG, among controls (n=29) and COVID-19 patients sampled in the sub-acute (n=15, denoted in red) or convalescent (n=36, denoted in orange) phase. **(B)** IgG levels expressed in relation to total IgG **(C)** Relationship between IgG to SARS-CoV-2 at the sub-acute time point and IgG to non-SARS HCoV.

### Application of the assay in COVID-19 cases followed longitudinally

Next, we used the assay to measure IgG antibodies to SARS-CoV-2 proteins in 17 patients hospitalized with a primary diagnosis of COVID-19 in which longitudinal blood draws were available. None of these patients had received convalescent plasma and the majority had severe disease as reflected by admission to the intensive care unit and a requirement of mechanical ventilation (online suppl. Table 1). Among these patients, the seroconversion rate was 88% for spike-RBD (15 of 17 patients) and 83% for nucleocapsid (14 of 17 patients), considering the whole duration of the study (Fig. 2). Of note, none of the patients who failed to seroconvert had samples available at time points later than 14 days post-symptom onset. IgG to both spike-RBD and nucleocapsid proteins generally occurred in parallel, were positive within 10-12 days after symptoms onset and decreased over time (Fig. 2 and online suppl. Fig. 4).

### Levels of IgG to SARS-CoV-2 in relation to non-SARS coronaviruses and reference antigens

We next sought to determine how levels of IgG to SARS-CoV-2 (spike-RBD) in the sub-acute (defined as day 14 to 28 post-symptom onset) and convalescent (post-hospitalization, median=70 days post-symptom onset) phase compared to baseline levels of IgG to the spike S1 protein of two non-SARS coronaviruses (HCoV). We focused on HCoV-229E and HCoV-OC43 as these two coronaviruses represent important causes of community upper respiratory viral infections [19]. We also assessed two reference antigens that were expected to be recognized by high levels of IgG. Tetanus toxoid represents an antigen to which most individuals are routinely immunized and galactose-α-1,3-galactose (α-Gal) is an oligosaccharide of non-primate mammals which is the target of abundant IgG in immunocompetent humans [20]. The results revealed that levels of IgG to SARS-CoV-2 in patients with severe COVID (in the sub-acute and convalescent phase) were higher than levels of IgG specific for HCoV-OC43 and HCoV-229E, and also tetanus toxoid or α-Gal (Mann-Whitney, *P*<0.01 for each comparison) (Fig. 3A). Total IgG levels trended higher in the COVID-19 patients but this did not achieve significance (Mann-Whitney, *P*=0.16). Responses to SARS-CoV-2 in the sub-acute phase represented nearly 1% of total IgG (Fig. 3B). Interestingly, IgG to α-Gal represented <0.1% of total IgG in both cases and controls, a level lower than previously reported [20]. Finally, this analysis allowed us to explore the effects of pre-existing immunity to HCoV on the magnitude of IgG to SARS-CoV-2 that developed in COVID-19 patients. Baseline levels of IgG to HCoV-229E and HCoV-OC43 spike S1 were paired with the highest value for IgG to SARS-CoV-2 that was observed in the sub-acute phase. Samples with the lowest levels of IgG to HCoV-229E and HCoV-OC43 appeared to have relatively higher levels of IgG to SARS-CoV-2, though this analysis was limited by sample size considerations (Fig. 3C).

## Discussion/Conclusion

The majority of serologic studies of COVID-19, and also commercial assays for measuring IgG to SARS-CoV-2, have been limited by a lack of quantitative information about anti-viral antibodies in standardized units. Here we have shown that the ImmunoCAP assay, which is well known for its strong performance characteristics for IgE measurement and is used in clinical and research labs across the globe, can be readily adapted for the quantitative measurement of IgG to SARS-CoV-2 proteins. The assay has high sensitivity and specificity and, unlike the majority of commercial assays, has a read-out in standardized units (µg/mL). A particular strength of the assay is that antibodies to multiple antigens, including novel antigens that have been linked to the solid phase by the biotin-streptavidin technique, can be run in parallel in a single run. Applying the assay, we have shown that peak levels of IgG to SARS-CoV-2 spike RBD and nucleocapsid can approach or exceed 100 µg/mL, equivalent to ∼1% of total IgG, in the sub-acute phase of infection before waning over time. This level significantly exceeded steady-state levels that were detected to major representative antigens, *ie* - IgG to tetanus toxoid, α-Gal and also to HCoV. The timing and trajectory of anti-SARS-CoV-2 IgG in our assay generally fit with prior reports, with levels peaking within 3 weeks of symptom onset and showing a significant decrease within 3 months [5, 21, 8, 22]. Although preliminary, our data also raise interesting questions about the role that pre-existing immunity to HCoV could play in affecting the development of IgG to SARS-CoV-2.

The current investigation of COVID-19 cases primarily included serum/plasma samples from those cases that were severe, as reflected by intensive care admission status and mechanical ventilation history. We acknowledge that the performance characteristics of the assay may differ when studying mild or asymptomatic COVID-19 cases. To date we have not compared the results from ImmunoCAP with results of neutralizing antibody assay. This is an important next step, however it has been convincingly reported that IgG to spike-RBD correlates with results using neutralization assays [23, 24]. Our data also suggests that non-specific binding (background) in this system is ∼1-3 µg/mL. This may be higher than background values observed in other assay systems, however, we have addressed this by accounting for binding to unconjugated streptavidin ImmunoCAPs for each sample. Acknowledging these limitations, the assay we have described has several strengths compared to many of the serological approaches currently being used in COVID-19 research and patient care. The ImmunoCAP-based assay we have described can readily be adapted to study IgG to SARS-CoV-2 and also a host of other antigens by using the streptavidin-biotin technique. The assay has good performance characteristics, has a quantitative read-out in standardized units and could be used by laboratories across the globe that routinely use the ImmunoCAP.

## Data Availability

Primary data is available on request from the corresponding author.

## Acknowledgement

William Petri, Pat Pramoonjago, Deborah Murphy, and colleagues affiliated with the University of Virginia COVID-19 Biorepository and clinical research teams. Jonas Lidholm at Thermo Fisher Scientific for helpful conversations about ImmunoCAP assay development. Christopher Moskaluk and Lindsay Bazydlo for facilitating access to samples from the clinical laboratory.

## Statement of Ethics

This study was approved by the Institutional Review Board for Health Sciences Research at the University of Virginia.

## Author Contributions

B.K. contributed to the study design, acquisition, analysis and writing of the paper. L.J.W. and M.D.S. contributed to the study design and sample acquisition and critically reviewed the manuscript. J.R.W, L.M.M, G.C., F.D., C.A.B, J.M.S., C.R., C.A.M., J.A.W., A.K., contributed to sample acquisition and critically reviewed the manuscript. T.A.E.P-M. and J.M.W. contributed to the study design, analysis, interpretation and writing of the paper. All authors have read and approved the final version of this paper.

