## Supplemental Files for "Quantitative measurement of IgG to SARS-CoV-2 proteins using ImmunoCAP"

**ONLINE SUPPLEMENT**

**Suppl. Table 1.** Characteristics of COVID-19 cases

| Characteristics | | COVID-19 Longitudinal cohort | COVID-19 Follow-up clinic* |
|---|---|---|---|
| N | | 17 | 36 |
| Age. Median (IQR) | | 65 (34-69) | 55 (48-66) |
| Sex | Male, n (%) | 11 (65%) | 21 (58%) |
| | Female, n (%) | 6 (35%) | 15 (42%) |
| Race/ ethnicity | Caucasian, n (%) | 6 (35%) | 6 (17%) |
| | Black, n (%) | 2 (12%) | 10 (28%) |
| | Hispanic, n (%) | 9 (53%) | 18 (50%) |
| Received IVIG or plasma product, n (%) | | 0 (0%) | 3 (8%) |
| Admitted to Intensive care unit, n (%) | | 15 (88%) | 35 (97%) |
| On mechanical ventilation, n (%) | | 13 (76%) | 32 (89%) |
| Survived hospitalization, n (%) | | 14 (82%) | 36 (100%) |
| Time between blood draw and symptom onset, days (IQR) | | 12 (7-17)** | 70 (56-79) |

*8 subjects were included in the longitudinal cohort and the follow-up clinic cohort.

**Days between first recorded symptom and first blood draw

**ONLINE SUPPLEMENT**

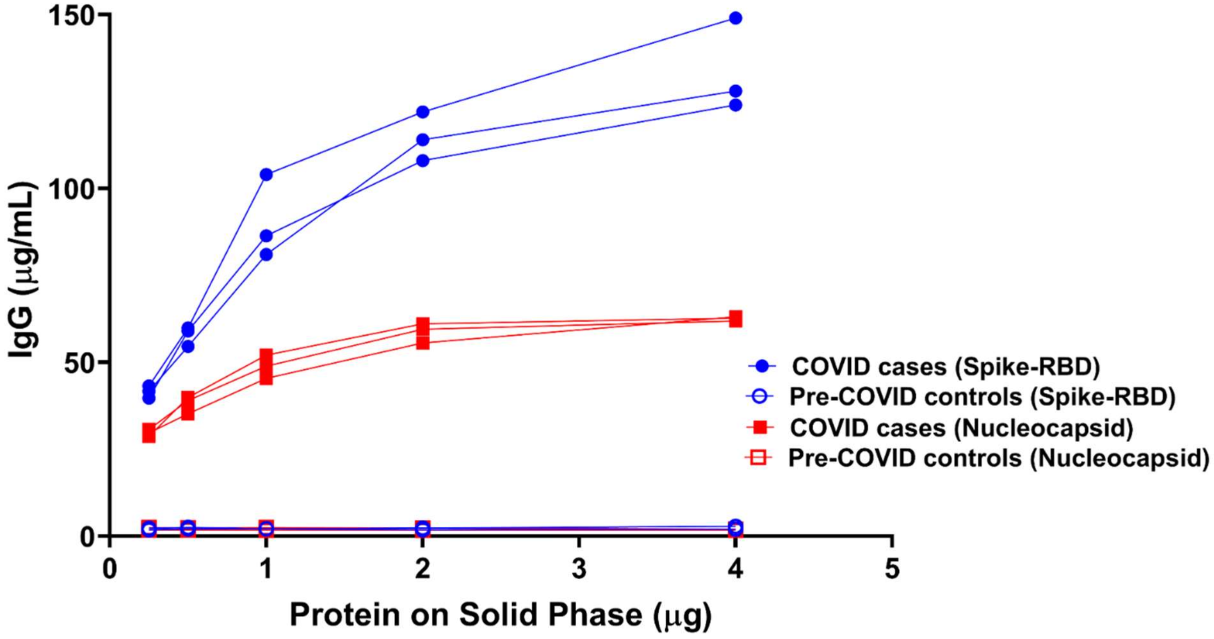


**Suppl. Fig. 1.** Measurement of IgG to SARS-CoV-2 spike-RBD and nucleocapsid using different amounts of viral antigen on the solid phase of the assay (N=3 patient samples).

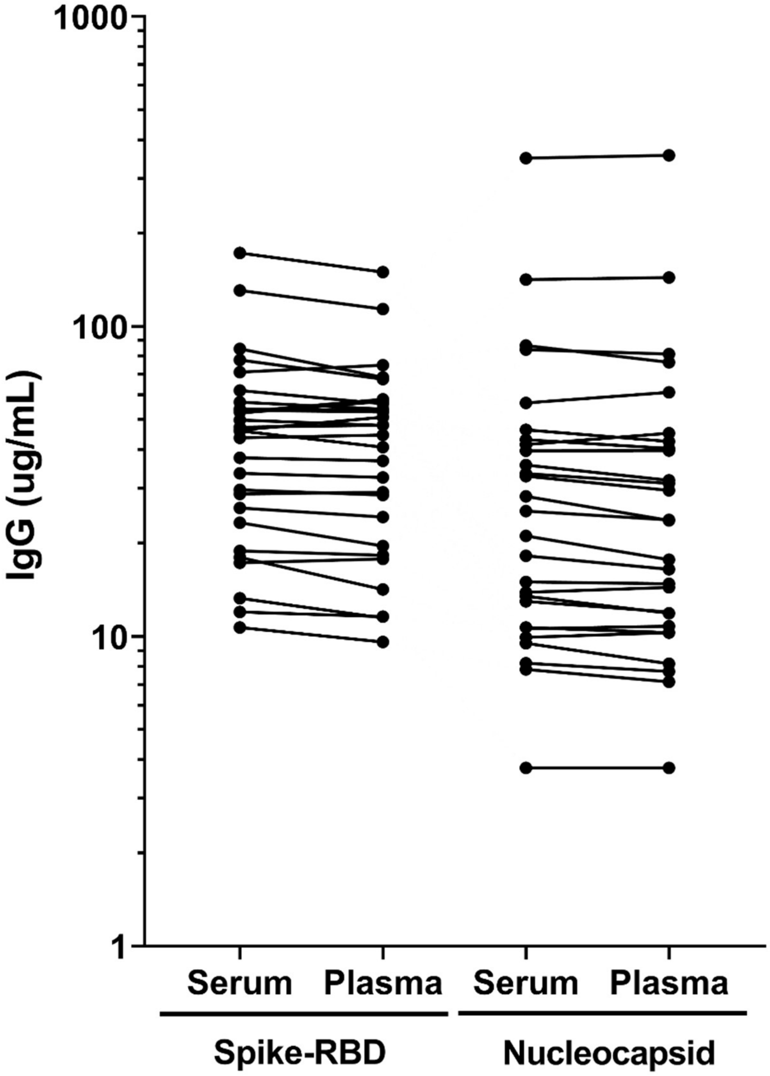


**Suppl. Fig. 2.** Comparison of IgG to SARS-CoV-2 spike-RBD and nucleocapsid using paired plasma and serum samples from COVID-19 patients (n=15).

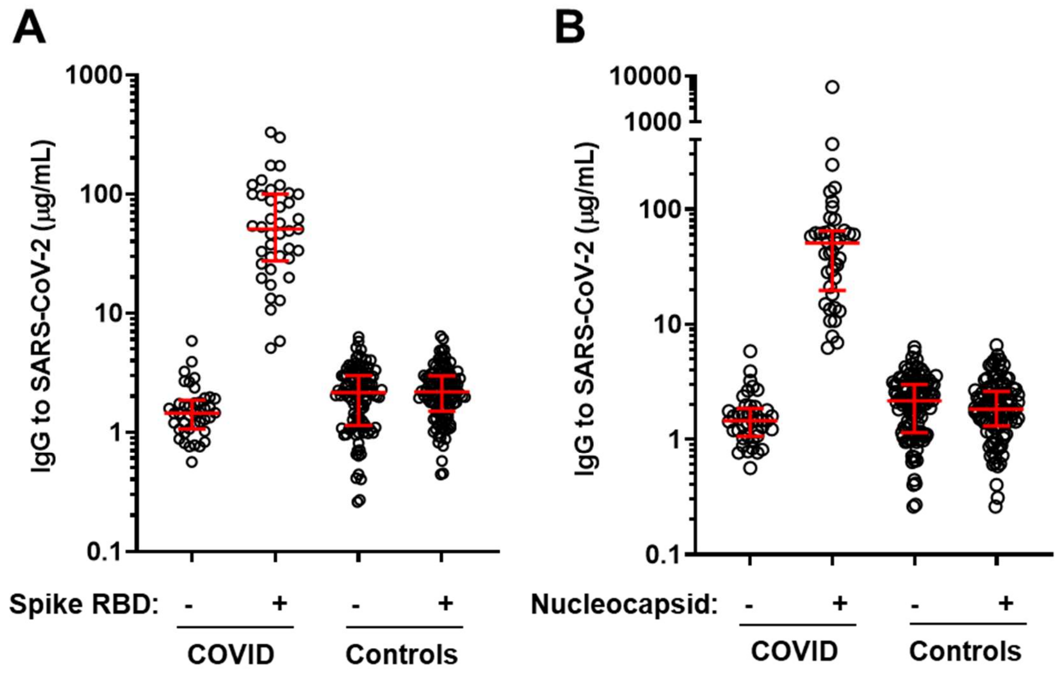


**Suppl. Fig. 3.** IgG to naked streptavidin ImmunoCAPs (-) or ImmunoCAPs carrying SARS-CoV-2 proteins (**A**, spike-RBD and **B**, nucleocapsid) on the solid-phase of samples from COVID-19 patients (n=41) or controls (n=109).

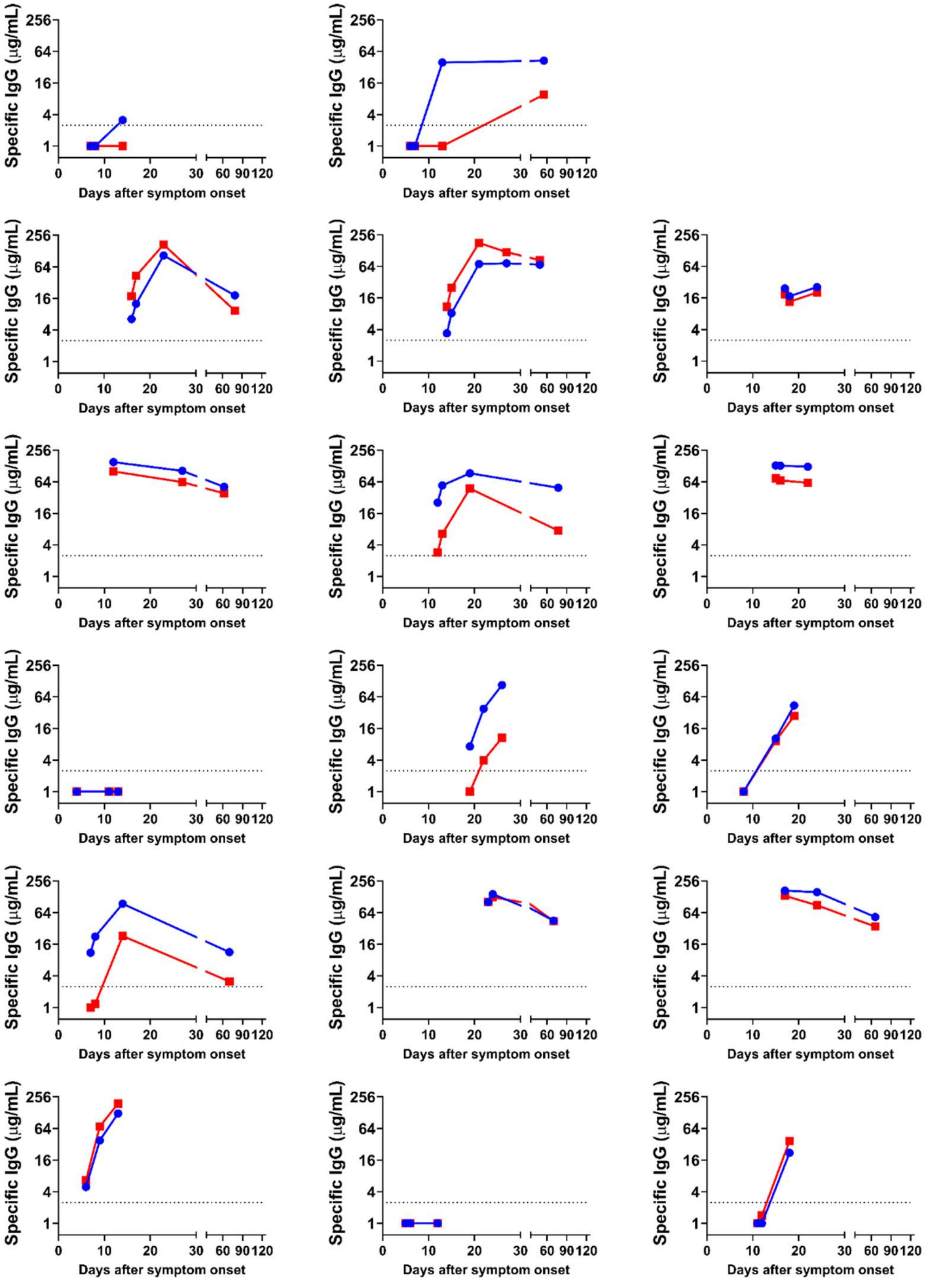


**Suppl. Fig. 4.** IgG to SARs-CoV-2 spike-RBD and nucleocapsid among 17 patients with severe COVID-19 who had longitudinal samples available.
